# Alcohol causes an increased risk of head and neck but not breast cancer in individuals from the UK Biobank study: A Mendelian randomisation analysis

**DOI:** 10.1101/19002832

**Authors:** Nathan Ingold, Hasnat A Amin, Fotios Drenos

## Abstract

Alcohol intake and the risk of various types of cancers have been previously correlated. Correlation though does not always mean that a causal relationship between the two is present. Excessive alcohol consumption is also correlated with other lifestyle factors and behaviours, such as smoking and increased adiposity, that also affect the risk of cancer and make the identification and estimation of the causal effect of alcohol on cancer difficult. Here, using individual level data for 322,193 individuals from the UK Biobank, we report the observational and causal effects of alcohol consumption on types of cancer previously suggested as correlated to alcohol. Alcohol was observationally associated with cancers of the lower digestive system, head and neck and breast cancer. No associations were observed when we considered those keeping alcohol consumption below the recommended threshold of 14 units/week. When Mendelian randomisation was used to assess the causal effect of alcohol on cancer, we found that increasing alcohol consumption, especially above the recommended level, was causal to head and neck cancers but not breast cancer. Our results where replicated using a two sample MR method and data from the much larger COGS genome wide analysis of breast cancer. We conclude that alcohol is causally related to head and neck cancers, especially cancer of larynx, but the observed association with breast cancer are likely due to confounding. The suggested threshold of 14 units/week appears suitable to manage the risk of cancer due to alcohol.

## INTRODUCTION

Harmful effects of excessive alcohol consumption have been suggested for various diseases, including liver cirrhosis, cancer and cardiovascular disease, as well as risk of injuries through accidents ^123^. In the case of the association of alcohol with cancer, both ethanol and the products of its metabolism from the cells are considered to be carcinogenic ^4^. A large body of work exists on the association between alcohol consumption and different types of cancer in model organisms and population samples ^5–7^. Recently, a meta-analysis of the cancer and alcohol observational literature produced a list of seven cancers, including cancers of the: oropharynx, larynx, oesophagus, liver, colon, rectum and female breast, that are associated with heavy alcohol consumption ^8^. These results have been widely publicized and currently form part of the official advice provided by the UK Chief Medical Officers’ Low Risk Drinking Guidelines. According to this consumption of alcohol should not exceed 14 units per week for both men and women ^9^.

There are multiple limitations of relying on observational associations to establish the causal role of alcohol on cancer and estimate its true effect on risk, especially when we are interested in either high or low levels of consumption. Alcohol consumption is associated with other cancer risk factors such as poorer diet high in fats and processed meat, obesity, and most importantly, smoking ^10,11^. Furthermore, measurement of alcohol consumption, in the vast majority of cases, relies on self-reported drinking habits. This requires the participants to recall how much they drink per day, week, or month period. Studies looking at the accuracy of self-reported alcohol consumption have found that under-reporting is common ^12^ and the inaccuracy of the provided information increases with increasing levels of consumption ^13^. Also, the longer the period required to recall, the less accurate the information provided is, with periods longer than a week considered unreliable ^14^. Although these limitations can be addressed through the use of randomised control trials, this is not an option in this case due to logistical and ethical problems. In such cases we can make use of the existing genetic variability as a natural randomised control study ^15^. Instead of randomly assigning individuals in cases and controls groups, we rely on genetic variation that affects alcohol consumption to identify those that are likely to consume more alcohol and those that are likely to consume less. As long as the chosen genetic variant is not associated with any of the possible confounders of the association between alcohol and cancer, we can use this variant as an unconfounded proxy of alcohol consumption to test and estimate the effect of the associated level of alcohol consumption on risk of cancer. This methodology is described as Mendelian Randomisation (MR) and has been previously used to estimate the causal effect of low to moderate alcohol consumption on cardiovascular risk using a genetic variant from the alcohol dehydrogenase 1B gene (*ADH1B*) ^3^. The *ADH1B* gene codes for the *ADH1B* enzyme metabolising alcohol into acetaldehyde. A single nucleotide polymorphism (SNP) found in the *ADH1B* gene, located at rs1229984, changes arginine to histidine in the encoded protein which results in increased levels of acetaldehyde after alcohol consumption and elicits a negative reaction to alcohol^16^.

Here using individual level data for 322,193 people from the UK Biobank study, we aimed to establish and estimate the causal effect of alcohol consumption on the risk of cancer types previously suggested as associated with alcohol consumption.

## METHODS

### Study population

The UK Biobank is a large population study including information and biological samples for approximately 500,000 individuals. These were recruited between 2006 and 2011. The 22 UK Biobank assessment centres throughout England, Wales and Scotland, collected baseline data from the participants in the form of questionnaires, physical and cognitive tests and blood and urine samples ^17^. The age range of the participants at the time of enrolment in the study was from 40 to 69 years, with a mean age of 56.5 years. Males represent 45.6% of the sample.

### Alcohol intake

Alcohol intake for each participant was measured through multiple questionnaires. Excessive drinking and drinking behaviour related problems were assessed through the mental health questionnaire. Consumption of alcoholic drinks in the past 24 hours was assessed through the diet questionnaire. We used average weekly intake data from the lifestyle and environment questionnaire, including information for both weekly and monthly consumption. These were split per alcoholic beverage. An example for the average weekly red wine intake question was: “In an average WEEK, how many glasses of RED wine would you drink? (There are six glasses in an average bottle)” ^18^. The mean answer to this question from 348,369 participants was 3.9 glasses per week. The total number of units consumed per week by each participants was calculated using information available from the NHS (see ^19^ and https://www.lanarkshirelinks.org.uk/wp-content/uploads/2015/12/HWL-ALCOHOL-KNOW-YOUR-LIMITS-SHEET.pdf). Participants who had indicated that they had reduced their alcohol consumption either due to their doctor’s advice or due to illness (UKB Field 2664) were excluded.

### Cancer information

Cancer diagnoses information for the UK Biobank participants was obtained through self-reported questionnaires and through linkage to existing cancer registries, the Medical Research Information Service for England and Wales and the Information Service Division for Scotland. The cancer registries include information from the early 1970’s acquired through a variety of sources, including hospitals and cancer treatment centres. More information can be found at http://biobank.ndph.ox.ac.uk/showcase/docs/CancerLinkage.pdf. The data are coded using the 10th revision of the International Classification of Diseases (ICD). All participants who had been diagnosed with a given cancer at any point were considered cases and those that had not been diagnosed with that cancer were considered controls. For this study, cancer outcomes for lip, oral cavity and pharynx (ICD 10: C00-C14), digestive organs (ICD10: C15-26), larynx (ICD 10: C32) and for female only cases of breast cancer (ICD 10: C50) were used. We also made larger categories of these by grouping C00-C14 with C32 as head and neck cancers, C15-C26 as gastro-intestinal cancers, C15-C16 and C22-C25 as upper digestive cancers, C17-C21 as lower digestive cancers, and all codes used in a category of any cancer. In terms of individual cancer types, we analysed only those that had at least 100 cases recorded in the study. In addition, acute myocardial infarction (MI) (ICD 10: I21) was used as a positive control with a known causal association from previous analysis.

### Genotyping

The rs1229984 variant was genotyped in 488,363 individuals as part of the two similar genome wide genotyping Affymetrix Axiom arrays used in the UK Biobank. Sample quality control metrics were provided by UK Biobank and were generated as described previously ^20^. Samples were excluded from the analysis if they were determined to be outliers for missingness or heterozygosity, and if they had any sex chromosome aneuploidies or differences between reported and genetically inferred sex. Samples which did not have a White British ancestry were also excluded from this analysis. A list of related individuals was provided by UK Biobank and one individual from each related pair was excluded at random. After filtering, a total of 333,774 participants remained in the sample genotyped for rs1229984.

### Statistical analysis

We used R 3.5.1 for analysing and plotting the results^21^, unless otherwise stated. For the observational part of the study, we regressed the amount of alcohol consumed per week against each of the outcomes considered using a robust logistic regression from the “robust” R package ^22^ adjusting for age and sex. The analysis was replicated per category of alcohol consumption grouped as below the recommended threshold (1-14 units or alcohol per week), or above the recommended threshold (<14 units or alcohol per week). We tested the association of the rs1229984 on alcohol consumption using a robust linear regression model including an additive and a dominance deviation component. Due to evidence of dominance, we repeated the analysis using a dominant minor allele coding for rs1229984 in all participants and in groups of alcohol consumption adjusting for age, sex, the first 4 principal components (PCs) for the genetic variability of the genome, and the genotyping array used. We tested the association of the genotype with each of the outcomes through a logistic regression adjusted for age, sex, the 4 PCs, and genotyping array used. The analysis was replicated per category of alcohol consumption including for those reporting 0 units of alcohol consumed per week. To assess the causal role of alcohol consumption on cancer risk, we used the ivprobit command in Stata 15 ^23^ with the robust option and the maximum likelihood estimator method, while adjusting for age, sex, PCs, and genotyping array used. The analysis was replicated for all other categories of alcohol consumption. To confirm our results we also used an external genome wide association (GWAS) of alcohol consumption that identified multiple genetic polymorphisms ^24^ that were used in a two sample MR through MR Base ^25^ with UK Biobank cancer data. For breast cancer, we also used an external outcome file from the publicly available COGS results ^26^. We accepted associations with a p-value < 0.05/15 = 0.003571 to adjust for the 15 different outcome measures tested. We also describe results with 95% confidence intervals (CIs) away from 0 or 1 accordingly but as probable associations.

## RESULTS

After filtering for poor genotyping and non-white British ethnic background and excluding those reporting changes in their drinking behaviour due to their doctor’s advice or due to illness, 322,193 individuals remained in the sample. The mean age of the sample was 56.8 years (Standard Deviation (SD) 8 years), with participants on average slightly overweight with a BMI of 27.4 kg/m^2^ (SD 4.7 kg/m^2^). The sample included 54% females, and 60% reported current or previous smoking. The average units consumed were 13 units/week (interquartile range 19 units/week). More than a quarter of the sample (28%) reported no alcohol intake per week, 38% were consuming alcohol below the recommended threshold of intake (1-14 units/week), and 34% above this (>14 units/week). The frequency of the minor T allele of the *ADH1B* rs1229984 SNP was 2.2%. There was no evidence of association between the variant and any of the common risk factors for cancer (ever smoked p-value = 0.197). Those carrying the minor rs1229984 allele were less likely to exceed the recommended level (p-value = 7.92×10^−132^) and more likely to report no average weekly alcohol consumption (p-value = 4.44×10^−104^). A summary of the overall sample separated per genotype is provided in Table 1.

**Table 1:** Characteristics of the sample used in the analysis. Summary for the entire sample and per genotype of the rs1229984 SNP is provided. There is no evidence that the rs1229984 SNP is associated with the selected classical risk factors for cancer. The distribution of alcohol consumption is not normally distributed and the standard deviations should be interpreted with caution.

| Genotype | N | % Female | Age (years) | BMI (kg/m<sup>2</sup>) | Ever smoked? (% Yes) | Alcohol<br>(units/wk) and<br>IQR | % Non-drinkers (0<br>units/wk) | % below recommended<br>threshold (1-14<br>units/wk) | % above recommended<br>threshold (>14<br>units/wk) |
| --- | --- | --- | --- | --- | --- | --- | --- | --- | --- |
| ALL | 322193 | 54.0 | 56.8 ±8.0 | 27.3 ±4.7 | 59.7 | 13.0 (19) | 0.28 | 0.38 | 0.34 |
| CC | 307752 | 54.1 | 56.8 ±8.0 | 27.3 ±4.7 | 59.6 | 13.2 (19) | 0.28 | 0.38 | 0.35 |
| CT | 14263 | 52.7 | 57.1 ±8.0 | 27.1 ±4.6 | 60.2 | 9.7 (14) | 0.36 | 0.39 | 0.25 |
| TT | 178 | 53.4 | 57.5 ±7.5 | 26.9 ±3.9 | 59.0 | 8.9 (15) | 0.45 | 0.29 | 0.26 |

Observational associations between alcohol and cancer revealed that increasing alcohol consumption was associated with an increasing risk of cancer, including those of the lower, but not upper, digestive system and head and neck cancers. In terms of specific locations, alcohol was associated with neoplasms of the tonsils, larynx, colon and rectum, as well as breast cancer. Alcohol appeared protective for acute MI, used as a control outcome with a known association. Odds ratios, 95% confidence intervals and p-values can be seen in Table 2. When the association between alcohol and cancer was tested in those drinking but within recommended intake limits, no evidence of association between level of alcohol consumption and cancer or acute MI was observed. In those exceeding the recommended level of alcohol, increasing intake was associated with cancers of the tongue, tonsils, larynx, oesophagus, colon, rectum, liver, and pancreas, as well as all broader categories tested (S Table 1). A probable association was present for stomach cancer but did not satisfy our multiple testing adjusted p-value threshold (S Table 1). No evidence of alcohol association with either acute MI or breast cancer were observed per category. No outcome had opposite direction of association per alcohol category. These results are graphically summarised in SFigure1.

**Table 2:**
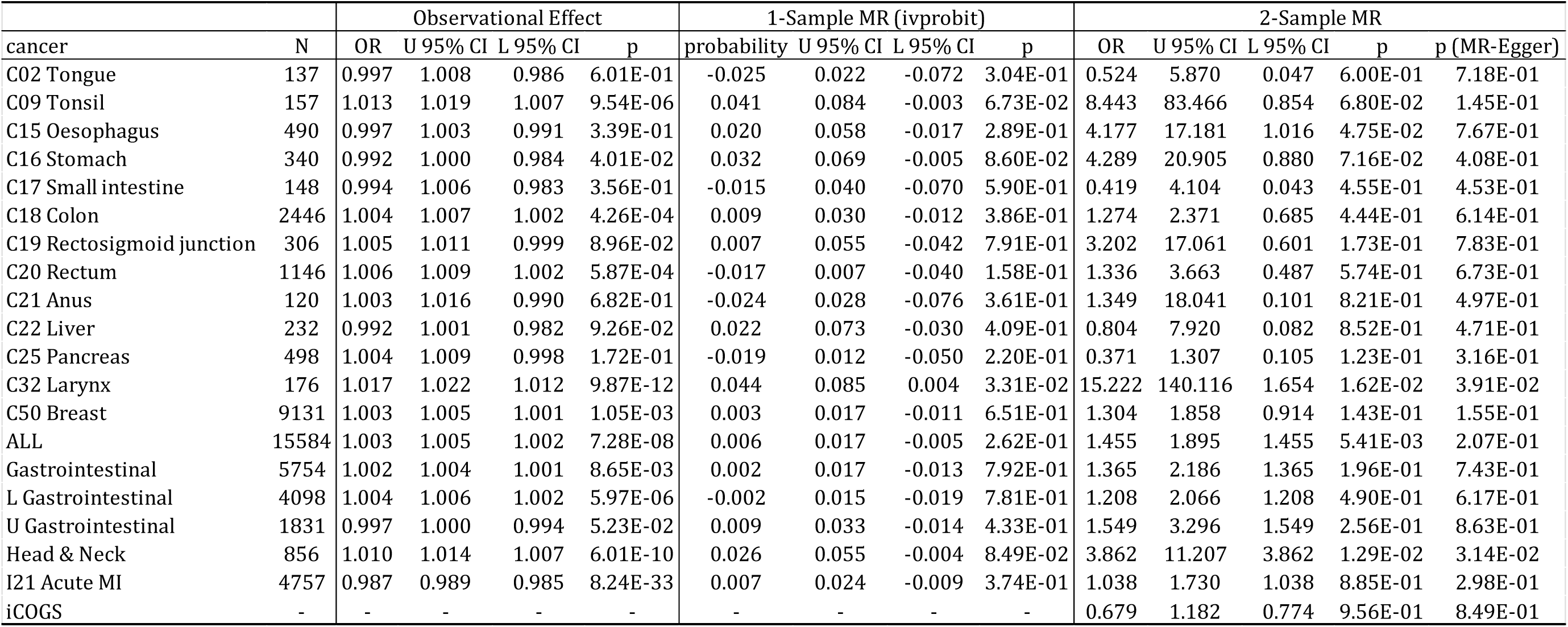
Results of observational, one and two sample MR analysis for the relationship between alcohol consumption in units/week and risk of cancer in all participants. U = upper; L = lower; CI = confidence interval; probability = ivprobit coefficient; MI = myocardial infarction. (An Excel version is available in the Supplementary Materials.)

| **Table 2** |  |  |  |  |  |  |  |  |  |  |  |  |  |  |
| --- | --- | --- | --- | --- | --- | --- | --- | --- | --- | --- | --- | --- | --- | --- |
|  |  | Observational Effect |  |  |  | 1-Sample MR (ivprobit) |  |  |  | 2-Sample MR |  |  |  |  |
| cancer | N | OR | U 95% CI | L 95% CI | p | probability | U 95% CI | L 95% CI | p | OR | U 95% CI | L 95% CI | p | p (MR-Egger) |
| C02 Tongue | 137 | 0.997 | 1.008 | 0.986 | 6.01E-01 | -0.025 | 0.022 | -0.072 | 3.04E-01 | 0.524 | 5.870 | 0.047 | 6.00E-01 | 7.18E-01 |
| C09 Tonsil | 157 | 1.013 | 1.019 | 1.007 | 9.54E-06 | 0.041 | 0.084 | -0.003 | 6.73E-02 | 8.443 | 83.466 | 0.854 | 6.80E-02 | 1.45E-01 |
| C15 Oesophagus | 490 | 0.997 | 1.003 | 0.991 | 3.39E-01 | 0.020 | 0.058 | -0.017 | 2.89E-01 | 4.177 | 17.181 | 1.016 | 4.75E-02 | 7.67E-01 |
| C16 Stomach | 340 | 0.992 | 1.000 | 0.984 | 4.01E-02 | 0.032 | 0.069 | -0.005 | 8.60E-02 | 4.289 | 20.905 | 0.880 | 7.16E-02 | 4.08E-01 |
| C17 Small intestine | 148 | 0.994 | 1.006 | 0.983 | 3.56E-01 | -0.015 | 0.040 | -0.070 | 5.90E-01 | 0.419 | 4.104 | 0.043 | 4.55E-01 | 4.53E-01 |
| C18 Colon | 2446 | 1.004 | 1.007 | 1.002 | 4.26E-04 | 0.009 | 0.030 | -0.012 | 3.86E-01 | 1.274 | 2.371 | 0.685 | 4.44E-01 | 6.14E-01 |
| C19 Rectosigmoid junction | 306 | 1.005 | 1.011 | 0.999 | 8.96E-02 | 0.007 | 0.055 | -0.042 | 7.91E-01 | 3.202 | 17.061 | 0.601 | 1.73E-01 | 7.83E-01 |
| C20 Rectum | 1146 | 1.006 | 1.009 | 1.002 | 5.87E-04 | -0.017 | 0.007 | -0.040 | 1.58E-01 | 1.336 | 3.663 | 0.487 | 5.74E-01 | 6.73E-01 |
| C21 Anus | 120 | 1.003 | 1.016 | 0.990 | 6.82E-01 | -0.024 | 0.028 | -0.076 | 3.61E-01 | 1.349 | 18.041 | 0.101 | 8.21E-01 | 4.97E-01 |
| C22 Liver | 232 | 0.992 | 1.001 | 0.982 | 9.26E-02 | 0.022 | 0.073 | -0.030 | 4.09E-01 | 0.804 | 7.920 | 0.082 | 8.52E-01 | 4.71E-01 |
| C25 Pancreas | 498 | 1.004 | 1.009 | 0.998 | 1.72E-01 | -0.019 | 0.012 | -0.050 | 2.20E-01 | 0.371 | 1.307 | 0.105 | 1.23E-01 | 3.16E-01 |
| C32 Larynx | 176 | 1.017 | 1.022 | 1.012 | 9.87E-12 | 0.044 | 0.085 | 0.004 | 3.31E-02 | 15.222 | 140.116 | 1.654 | 1.62E-02 | 3.91E-02 |
| C50 Breast | 9131 | 1.003 | 1.005 | 1.001 | 1.05E-03 | 0.003 | 0.017 | -0.011 | 6.51E-01 | 1.304 | 1.858 | 0.914 | 1.43E-01 | 1.55E-01 |
| ALL | 15584 | 1.003 | 1.005 | 1.002 | 7.28E-08 | 0.006 | 0.017 | -0.005 | 2.62E-01 | 1.455 | 1.895 | 1.455 | 5.41E-03 | 2.07E-01 |
| Gastrointestinal | 5754 | 1.002 | 1.004 | 1.001 | 8.65E-03 | 0.002 | 0.017 | -0.013 | 7.92E-01 | 1.365 | 2.186 | 1.365 | 1.96E-01 | 7.43E-01 |
| L Gastrointestinal | 4098 | 1.004 | 1.006 | 1.002 | 5.97E-06 | -0.002 | 0.015 | -0.019 | 7.81E-01 | 1.208 | 2.066 | 1.208 | 4.90E-01 | 6.17E-01 |
| U Gastrointestinal | 1831 | 0.997 | 1.000 | 0.994 | 5.23E-02 | 0.009 | 0.033 | -0.014 | 4.33E-01 | 1.549 | 3.296 | 1.549 | 2.56E-01 | 8.63E-01 |
| Head & Neck | 856 | 1.010 | 1.014 | 1.007 | 6.01E-10 | 0.026 | 0.055 | -0.004 | 8.49E-02 | 3.862 | 11.207 | 3.862 | 1.29E-02 | 3.14E-02 |
| I21 Acute MI | 4757 | 0.987 | 0.989 | 0.985 | 8.24E-33 | 0.007 | 0.024 | -0.009 | 3.74E-01 | 1.038 | 1.730 | 1.038 | 8.85E-01 | 2.98E-01 |
| iCOGS | - | - | - | - | - | - | - | - | - | 0.679 | 1.182 | 0.774 | 9.56E-01 | 8.49E-01 |

The rs1229984 *ADH1B* variant showed evidence of dominance of the minor allele (p = 0.0236). Using a dominant model, the variant was strongly associated with alcohol intake (F-statistic 692.2, p-value 5.62×10^−106^) in the sample. Carriers of the minor T allele were consuming on average 2.1 units/week less that the homozygotes of the common allele. The association of the genetic variant with alcohol was also evident for both of those drinking below (F-statistic = 47.4, p-value = 2.63×10^−13^) and above (F-statistic = 69.9, p-value = 1.20×10^−6^) the maximum recommended 14 units/week, with a difference of 0.35 and 0.87 units/week less respectively. A genetic variant, or allele score, with an F-statistic > 10 for its association with the exposure is considered a strong instrument for use in MR. Testing the association between the variant and the cancer outcomes though, showed no evidence of association between the rs1229984 *ADH1B* variant and any of the cancer outcomes tested in the full sample, those reporting alcohol consumption of 1 to 14 units/week, or those drinking more than 14 units/week (STable 2). For those reporting no alcohol consumption per week, a probable association suggesting an increase of risk of rectal cancer with the minor allele was observed, but this is well within our expected false positives range (STable 2). In those exceeding the recommended limit of alcohol consumption, we identified a protective effect of the minor allele of the genetic variant reducing alcohol use and acute MI (Odds ratio 0.61, CIs 0.44-0.85, p-value = 2.95×10^−3^) (STable 2).

The instrumental variable regression estimating the causal effect of alcohol consumption on the risk of cancer in all individuals showed no evidence of a causal effect at our multiple testing adjusted p-value threshold, though a probable association was seen with the risk of larynx cancer (probit coefficient 0.044; CIs 0.004-0.085) (Table 2). No evidence of a causal association between alcohol consumption and cancer risk for those reporting that they drink but on average less than 14 units/week was observed. Those exceeding the suggested limit however showed evidence of causal association between alcohol and head and neck cancer (0.053; CIs 0.033-0.073). Probable associations were also seen with an increase of the risk of larynx cancer (0.049; CIs 0.001-0.097), and a decrease of the risk of tongue cancer (−0.47; CIs -0.082 --0.012) with alcohol, though their respective p-values do not reach our multiple testing adjusted threshold (STable 3). For risk of MI we identified evidence of association with alcohol consumption in those drinking more than the recommended maximum (0.048 CIs 0.033-0.062) but not for those drinking below it or in the sample overall (STable 3). The results are visually summarised in Figure 1.

**Figure 1.**
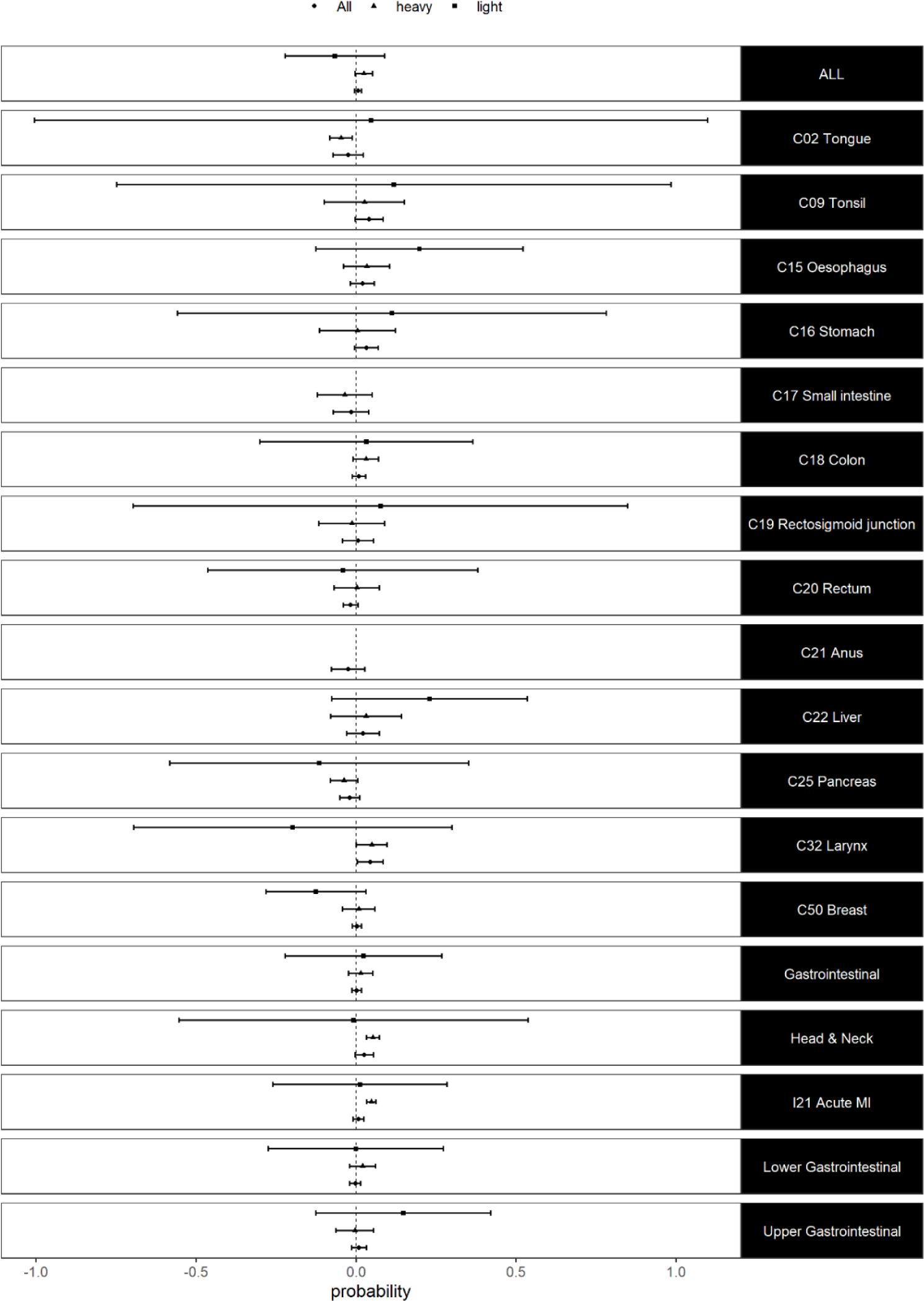
Results of 1-Sample MR (ivprobit) analysis for the relationship between alcohol consumption in units/week and cancer in light (≤14 units) and heavy (>14 units) drinkers and in all participants

The two sample MR using the recently published polygenic associations with alcohol consumption ^24^ did not fundamentally change our results (Table 2). The biggest difference was the probable causal association of any cancer category with alcohol consumption (OR 1.45, CIs 1.12-1.89), though MR Egger results did not support this effect. Alcohol had a probable causal association to head and neck cancer for both the inverse variance weighted (OR 3.86, CIs 1.33-11.21) and MR Egger method. Similarly, cancer of the larynx had again evidence of a probable causal association not reaching our multiple testing adjusted p-value (OR 15.22, CIs 1.65-140.12). Cancer of the oesophagus also showed probable evidence of a causal association (ORs 4.18, CIs 1.02-17.18). Alcohol consumption was not associated with the risk of breast cancer in either of the analyses we performed using events in UK Biobank or when we used outcome associations from the largest breast cancer GWAS available from COGS ^26^ (OR 0.96, CIs 0.77-1.18) (Table 2 & SFigure 2).

## DISCUSSION

We set out to identify and estimate the causal effect of self-reported weekly alcohol consumption on the risk of cancer in the parts of the body in direct contact with alcohol during digestion. We also included breast cancer as a common cancer correlated to alcohol consumption and acute MI as a control outcome for which the causal effect of alcohol was previously well established ^3,27^. We found observational associations between weekly alcohol consumption and cancers of the digestive system. The genetic variant chosen as an instrument for alcohol consumption was strongly associated with the exposure but not with the cancer outcomes. In those exceeding the recommended maximum of 14 units/week, alcohol was causally associated with the risk of head and neck cancer. Only limited evidence was available for the causal effect of alcohol to specific cancers and these were focused on cancer of the larynx.

In terms of the observational associations between alcohol consumption and risk of cancer, our result correspond well to what has been previously reported in a meta-analysis of 572 studies ^8^. We confirmed the association of alcohol consumption with an increasing occurrence of cancers in the area of the pharynx (specifically tonsils), as well as colorectal, larynx and breast cancer. We did not replicate the association with oesophageal or liver cancers, except for those consuming more than the recommended maximum of 14 units/week, which also showed an increasing number of cancers of the oral cavity and pharynx, colorectal, pancreas and larynx. Small intestine was not affected by alcohol in any of the categories tested, in both our work and the published meta-analysis. For individuals drinking less than the recommended threshold we did not identify any associations of alcohol consumption with cancer which is in accordance with the published data, except for oral cavity and pharynx ^8,28^. A J or U shaped correlation has been suggested between alcohol and risk of cardiovascular disease ^29,30^. Here using acute MI we found a protective effect of alcohol in the overall sample that attenuated in magnitude with increasing category of alcohol consumption in accordance with the idea of a non-linear relationship.

The rs1229984 variant we selected as the genetic instrument, based on other published MR work ^3,27,25^, was strongly associated with weekly alcohol consumption in our sample. The variant also had other characteristics of a good instrument, such as no evidence of an association of the genetic variant with any of the probable confounders or with any of the cancer types tested in those reporting no alcohol consumption per week. Though, the latter should be considered with caution, since an individual’s report of no weekly alcohol consumption does not rule out the occasional drink. We did not find any associations between the variant and the cancer types tested in the overall sample. The lack of association of the specific variant with stomach cancer agrees with other published data ^22,31,32^, though other alcohol associated SNPs have been reported as associated with the outcome. The rs1229984 variant though has been previously associated with aerodigestive cancers, including oral and pharynx, larynx and oesophagus cancers ^33,34,35^. Despite not identifying the reported associations, our estimated effects per unit/week were very close to what was reported earlier in a sample of 3,800 cases and 5,200 controls ^33^, suggesting that more cancer cases were required to identify some of the associations present. There is some uncertainty for the association of the specific variants with colorectal cancers with some studies reporting an association ^36^ and others reporting no effect ^37^. We did not find an association of the variant with any of the lower digestive system cancers. Although the association of the variant used has been previously shown ^3^ and confirmed in a different population ^27^ we were only able to replicate the result in those reporting an alcohol intake higher than the recommended threshold.

Although our variant associations failed to identify causal effects reported in the literature, the instrumental variable regression strongly suggested that alcohol consumption higher than 14 units/week was causal to the risk of head and neck cancer and MI. The effect on head and neck cancer appeared to be, at least, partially attributed to an increase risk of cancer of the larynx. These results correspond well to the published associations of the genetic variant, described earlier ^33,34,35^, as well as studies showing that laryngeal cancers are reversible following alcohol cessation ^38^. The observed non-significant increase in the risk of cancer for those consuming alcohol in light and moderate quantities below the recommended threshold has been previously reported ^28^ though conflicting observational results are also available, especially for breast cancer. We did not identify any evidence of causal association between alcohol consumption and breast cancer, either for the 3,170 UK Biobank or the 122,977 COGS recorded events, irrespective of genetic instrument used. This suggests that the relationship between alcohol and breast cancer ^39^ is at least severely confounded. Smoking, poor diet and obesity are possible confounders that can also affect the suggested mechanisms for the link between alcohol and breast cancer ^40^.

Limitations exist in our study and they can explain some of the inconsistences we see with published results. The main issue with our data is the lack of statistical power due the relatively small number of recorded events. Although the study population is very large, reaching the 500,000 individuals, it includes people that are more affluent, better educated and in better health than the underlying population of the UK at this age range ^41^. This is evident for both the number of cancer events, which are 10-20% lower than expected, and alcohol consumption, with UK Biobank participants less likely to be alcohol abstainers but also less likely to drink alcohol every day compared to the general population ^41^. For liver cancer, where alcohol is a well-known major factor for primary hepatocellular cancers ^42^, our results should be interpreted as not covering the directly alcohol associated carcinomas. This is due to our removal from the data of those that changed their alcohol consumption following their doctor’s advice or illness, likely to include these with alcohol related liver disease, and due to that the vast majority of liver cancer cases attributed to cancer cells traveling to the liver from other parts of the body, instead of being primary liver tumours.

To summarise, we showed that alcohol consumption causes cancer of the head and neck and acute myocardial infarction in those exceeding the recommend threshold of 14 units/week. No causal effect of alcohol on cancer was found in those drinking below the threshold. No causal effect of alcohol was observed between alcohol consumption and breast cancer in women, in either the individual level data available here or previously published summary data, suggesting that the observational associations are more likely confounded. Our results contribute to the accumulating evidence for the danger posed from excessive alcohol use for some types of cancer, but not others, and the introduction of the lower 14 units/week threshold as a safe level of alcohol consumption in terms of cancer risk management.

## Data Availability

This research has been conducted using the UK Biobank Resource under project 44566 (https://www.ukbiobank.ac.uk/2018/12/genetic-and-non-genetic-factors-able-to-predict-and-modify-the-risk-of-different-types-of-cancer/). All bona fide researchers can apply to use the UK Biobank resource for health related research that is in the public interest.

---

**Supplementary Table 1**

Results of observational analysis for the relationship between alcohol consumption in units/week and cancer in light (≤14 units) and heavy (>14 units) drinkers.

**Supplementary Table 2**

Results of association analysis for the relationship between rs1229984 and cancer in non-drinkers, in light (≤14 units) and heavy (>14 units) drinkers, and in all participants. Values marked as FAIL indicate that no carriers were present in the cases for a given cancer so the results from the analysis cannot be interpreted for that specific cancer.

**Supplementary Table 3**

Results of 1-Sample MR (ivprobit) analysis for the relationship between alcohol consumption in units/week and cancer in light (≤14 units) and heavy (>14 units) drinkers. Values marked as FAIL indicate that no carriers were present in the cases for a given cancer so the results from the analysis cannot be interpreted for that specific cancer.

**Supplementary Figure 1**

Results of observational analysis for the relationship between alcohol consumption in units/week and cancer in light (≤14 units) and heavy (>14 units) drinkers and in all participants

**Supplementary Figure 2**

Results of 2-Sample MR analysis for the relationship between alcohol consumption in units/week and cancer in all participants

